# Predictive markers of response and toxicity to tarlatamab in patients with extensive stage small cell lung cancer: a multi-institutional real-world analysis

**DOI:** 10.64898/2026.10.02.26364624

**Authors:** Graeme Fenton, Wanru Guo, Daniel L. Hess, Annie L. Zhang, Curtis Tatsuoka, Alexis L. Green, Michelle Sittig, Ranee Mehra, Alexandra Simms, Afshin Dowlati, Taofeek K. Owonikoko, Katherine Scilla, Melinda Hsu, Laura Alder, Samuel Rosner

## Abstract

**Purpose:** Tarlatamab, a bispecific T-cell engager, has generated favorable response and survival outcomes in relapsed small cell lung cancer (SCLC). Factors influencing tarlatamab-associated treatment outcomes and toxicity have yet to be established. We performed a multivariate analysis to identify biomarkers associated with cytokine release syndrome (CRS) and immune effector cell associated neurotoxicity syndrome (ICANS) and integrated these variables into a novel risk stratification tool to identify patients at high risk of clinically significant toxicity.

**Patients and Methods:** Logistic and ordinal regression analyses evaluated the predictive value of variables contributing to the presence and severity, respectively, of CRS and ICANS in 115 patients with ES-SCLC receiving tarlatamab. Selected variables were then tuned using 5-fold cross-validation and incorporated into novel elastic net-based prediction models.

**Results:** Rates of observed CRS, ICANS, and dysgeusia were 46%, 28%, and 47%, respectively. ORR (overall response rate) and DCR (disease control rate) were 44% and 58.2%, while 6-month PFS (progression free survival) and OS (overall survival) rates were 30.4% and 54.9%, respectively. Excluding patients on tarlatamab <30 days, presence of dysgeusia was associated with a lower rate of disease progression (HR 0.44, p=0.00691) and death (HR 0.34, p=0.0145). The presence of, increased size and number of extracranial metastases as well as elevated baseline LDH were significantly associated with increased likelihood of CRS. Increased ECOG performance status, increased volume of brain metastases, development of CRS, and elevated baseline LDH were all associated with increased risk of developing ICANS. Our cross-validated risk model successfully predicted CRS grade ≥2 and ICANS events with 90.9% and 75% sensitivity, respectively.

**Conclusions:** This exploratory analysis identified several predictive biomarkers of toxicity that successfully stratified low and high-risk populations as part of an internally validated novel risk scoring tool.

**Statement of Translational Relevance:** Tarlatamab, a bispecfic T-cell engager (BiTE), is a novel immunotherapeutic agent for the treatment of relapsed extensive-stage small cell lung cancer (ES-SCLC). Cytokine release syndrome (CRS) and immune effector cell-associated neurotoxicity syndrome (ICANS) are well-documented side effects of tarlatamab, however factors influencing treatment outcomes and toxicity have yet to be fully established. We performed a retrospective multivariate analysis of SCLC patients receiving tarlatamab to assess the association between baseline patient characteristics and the likelihood of developing treatment response and toxicity. Dysgeusia was identified as a predictive indicator of positive response, while CRS and ICANS had no association with favorable treatment outcome. Several clinical variables were found to be associated with CRS and ICANS development and were incorporated into a risk analysis that successfully stratified patients based on likelihood of developing toxicity. This risk stratification algorithm represents a novel tool to help inform patient monitoring needs, treatment planning, and prophylaxis considerations, though further prospective assessment and validation is needed.

## Introduction

Small cell lung cancer (SCLC) accounts for 15% of lung cancer cases worldwide and is characterized by its aggressive biology, poor prognosis and early metastatic potential (1). While SCLC initially responds well to first line treatment, relapse is common, and subsequent treatment options have historically provided limited clinical benefit (2,3).

Following FDA approval in 2024, tarlatamab, a bispecfic T-cell engager (BiTE) targeting delta-like ligand 3 (DLL3) on tumor cells and CD3 on T-cells, has generated impressive treatment responses and sustained disease control in patients with relapsed SCLC (4). Recent retrospective analyses have reinforced this clinical benefit in real-world patient populations, while underscoring the unique toxicity profile of tarlatamab, including cytokine release syndrome (CRS), immune effector cell associated neurotoxicity syndrome (ICANS), and dysgeusia (5–7). While predictive markers of toxicity associated with T-cell engager therapies and immunotherapies used in the treatment of hematologic malignancies have been identified, markers associated with tarlatamab treatment response and toxicity have yet to be established (8–10).

With tarlatamab increasingly utilized in standard of care management for SCLC, the ability to effectively stratify patients based on predicted risk of toxicity has important clinical and financial implications in terms of mitigating risk and resource allocation. Due to the unique adverse events associated with tarlatamab, previous FDA regulations stipulated a minimum 22-hour inpatient observation period with the first two doses of tarlatamab (11). These monitoring requirements have limited access to tarlatamab, particularly in community cancer centers that lack inpatient observation capabilities. As of September 2026, updated FDA regulations now recommend an abbreviated 6 to 8 hour monitoring period for the first two tarlatamab infusions with in-person follow-up, including vital signs, the day after infusion. While these changes may help to expand access to tarlatamab and reduce associated healthcare resource utilization, the increased rate of CRS and ICANS observed in real-world populations suggests select patients, with increased risk, may still benefit from inpatient monitoring. As such, the ability to risk stratify patients and distinguish high risk candidates requiring increased monitoring may allow centers to better marshal resources to mitigate such toxicity, including potential targeted prophylactic strategies.

Herein, we present the largest multivariate analysis of real world tarlatamab data to date and identify several biomarkers of treatment response and toxicity. The subsequent risk model utilizing these biomarkers is, to our knowledge, the first such stratification tool specifically designed to identify patients at risk of developing clinically relevant toxicities while undergoing tarlatamab treatment.

## Materials and Methods

### Study Design and Patients

We performed a retrospective, multivariate analysis of patients with SCLC receiving tarlatamab at the University of Maryland Medical Center, Duke University, and Case Western Reserve Hospital. All patients who received at least one dose of tarlatamab at these institutions between July 2024 and October 2025 were eligible for inclusion. Data was collected under Institutional Review Board approved protocols at each respective center. Owing to the retrospective nature of the study, the requirement for written informed consent was waived.

Patients received tarlatamab per FDA label in 28-day cycles, with inpatient observation for at least 22 hours following tarlatamab infusion on day 1 and day 8 of cycle 1. No patients received prophylactic steroids or tocilizumab prior to treatment initiation. Pending tolerability during inpatient monitoring, subsequent tarlatamab infusions were administered outpatient utilizing recommended observation periods as per drug label. Baseline biomarkers — LDH, ferritin, uric acid, CRP — were collected on cycle 1 day 1 (C1D1) prior to infusion of tarlatamab.

### Study Endpoints

Treatment response was determined using clinical interpretation of radiological disease assessment from patients with available post-treatment imaging. CRS and ICANS were reported using American Society for Transplantation and Cellular Therapy (ASTCT) consensus guidelines(12). Dysgeusia was graded using Common Terminology Criteria for Adverse Events (CTCAE) version 5.0. CRS, ICANS, dysgeusia, thrombocytopenia and neutropenia were labeled side effects of special interest (SESI) for this analysis.

### Statistical Considerations

Univariate analyses assessing CRS and ICANS occurrence, ORR (overall response rate), and DCR (disease control rate) were performed using Chi – square/ Fisher’s exact tests. Overall survival (OS) and progression-free survival (PFS) univariate analyses were performed using log rank tests. When assessing dysgeusia and its association with outcomes, patients who received tarlatamab <30 days were excluded, given the known timing of this specific side effect.

Variables with associated p-values less than 0.05 identified through univariate analysis were subsequently included in multivariate analyses. Multivariate logistic regressions adjusted for age, sex and ECOG performance status were conducted for CRS, ICANS, ORR and DCR. Cox proportional hazards models were used to test significance of predictors on OS and PFS. The proportional hazards assumptions were assessed by Schoenfeld residual analyses. Statistically significant variables identified in the multivariate logistic regressions were then subjected to variable selection in parsimonious elastic net-based prediction models for both CRS and ICANS. For the purposes of risk stratification, clinically significant toxicity was considered development of CRS grade ≥2 and/or any grade ICANS. The parameters for each EN model (*λ*, *α*) were tuned using 5-fold cross-validation to maximize area under the curve (AUC) (Supplemental Table 1). Model performance was reported using pooled out-of-fold cross-validation (OOF CV) predictions to determine an aggregate prediction model, with thresholds selected by Youden’s index. The thresholds for meeting high risk criteria for CRS and ICANS were 0.252 and 0.241, respectively, chosen by cross-validation. Final full-data metrics were determined after refitting the selected model on the complete analytic cohort. All statistical analysis were conducted in R (version 4.6.1).

## Results

### Patient Characteristics

A total of 115 patients received at least one dose of tarlatamab and were included in the retrospective analysis (Table 1). Median age at C1D1 was 67 years (range 43-86), 56% were female (n = 64), and 79% were white (n = 91). A majority of patients had a current (20%, n = 23) or former (73%, n = 84) tobacco use history. 23% (n = 27) of patients had an Eastern Cooperative Oncology Group (ECOG) performance status score of ≥ 2, with median prior lines of therapy of 2 (range 1–5). 42% (n = 48) and 52% (n = 60) of patients had liver metastases and brain metastases at treatment start, respectively, with 23% (n=27) patients having untreated brain metastases at treatment start.

**Table 1.** Baseline Patient Demographics of the Study Population.

**A.**
|  | **n (%), n = 115** |
| --- | --- |
| **Median age (range) - years** | 67 (43-86) |
| **Sex - no. (%)** |  |
| **Male** | 51 (44.3) |
| **Female** | 64 (55.7) |
| **Race or ethnic group - no. (%)** |  |
| **Black** | 18 (15.7) |
| **Native American** | 3 (2.6) |
| **Non-Hispanic or Latino White** | 91 (79.1) |
| **Unknown** | 3 (2.6) |
| **Smoking history** |  |
| **Never** | 8 (7.0) |
| **Current** | 23 (20.0) |
| **Former** | 84 (73.0) |
| **ECOG** |  |
| **0** | 13 (11.3) |
| **1** | 75 (65.2) |
| **2** | 32 (20.0) |
| **3** | 4 (3.5) |
| **No. of previous lines of therapy - no. (%)** |  |
| **1** | 58 (50.4) |
| **2** | 42 (36.5) |
| **3** | 10 (8.7) |
| **>3** | 5 (4.4) |
| **Median no. of previous lines of therapy (range)** | 2 (1-5) |
| **Liver metastases at tarlatamab start** | 48 (41.7) |
| **Sum of liver lesion diameters > 100 mm** | 18 (15.7) |
| **Brain metastases at tarlatamab start** | 60 (52.2) |

### Side Effects of Special Interest

Of the 115 patients, 93 (80.9%) experienced a SESI after treatment initiation (Table 2). CRS was observed in 53 patients (46.1%): grade 1 in 22.6% (n= 26), grade 2 in 19.1% (n=22), grade 3 in 3.4% (n=4), grade 4 in 1.0% (n=1). Grade 4 CRS was observed in a patient requiring urgent initiation of tarlatamab given extensive disease progression complicated by new pulmonary embolism. ICANS was observed in 32 patients (27.8%): grade 1 in 12.2% (n= 14), grade 2 in 13.0% (n=15), grade 3 in 2.6% (n=3). All Grade 3 ICANS events were observed in patients with known intracranial disease. No grade 4 ICANS was observed. A majority of CRS (75.5%) and ICANS (65.6%) events occurred within 24 hours of C1D1. CRS and ICANS had a median onset time following start of C1D1 tarlatamab infusion of 12 and 20 hours, respectively. Median duration of both CRS and ICANS to symptomatic resolution was less than 24 hours.

**Table 2.** Immune Related Adverse Events on Tarlatamab.

|  |  | **n (%), n = 115** |
| --- | --- | --- |
| **Any Adverse Event** |  | **93 (80.9)** |
| **CRS** |  |  |
| Overall |  | 53 (46.1) |
| Grade 1 |  | 26 (22.6) |
| Grade 2 |  | 22 (19.1) |
| Grade 3 |  | 4 (3.4) |
| Grade 4 |  | 1 (1.0) |
| Median time to onset (range) - days |  | 0 (0-58) |
| Median time to onset after most recent infusion (range) - hours |  | 12 (7-36) |
| Duration of symptoms (range) - days |  | 1 (1-4) |
| Received Tocilizumab |  | 21 (18.3) |
| Average number of Tocilizumab doses |  | 1.3 |
| **ICANS** |  |  |
| Overall |  | 32 (27.8) |
| Grade 1 |  | 14 (12.2) |
| Grade 2 |  | 15 (13.0) |
| Grade 3 |  | 3 (2.6) |
| Median time to onset (range) - days |  | 1 (0-133) |
| Median time to onset after most recent infusion (range) - hours |  | 20 (6-72) |
| Received Dexamethasone |  | 27 (23.5) |
| With brain metastases at treatment initiation |  | 23 (20.0) |
| **Neutropenia** |  |  |
| Overall |  | 11 (9.6) |
| Grade 1 |  | 5 (4.4) |
| Grade 2 |  | 3 (2.6) |
| Grade 3 |  | 3 (2.6) |
| **Dysgeusia** |  |  |
| Overall |  | 54 (47.0) |
| Grade 1 |  | 12 (10.4) |
| Grade 2 |  | 42 (36.5) |
| Median time to onset (range) - days |  | 28 (2-265) |
| **Thrombocytopenia** |  |  |
| Overall |  | 23 (20.0) |
| Grade 1 |  | 15 (13.0) |
| Grade 2 |  | 2 (1.7) |
| Grade 3 |  | 4 (3.5) |
| Grade 4 |  | 2 (1.7) |

Dysgeusia was observed in 54 patients (47.0%), with 42 patients (36.5%) developing grade 2 events. Neutropenia and thrombocytopenia were observed in 10 patients (8.7%) and 23 patients (20.0%), respectively.

### General Efficacy

Ninety-one patients, with available post-treatment imaging, were included in our response analysis. Of the patients, 40 (44.0%) experienced clinically assessed objective response, with eight patients (8.8%) achieving complete response (CR). DCR was 58.2%. After a median follow-up time of 3.6 months, median OS and PFS was 9.1 months (95% CI: 5.5 – not reached) and 2.9 months (95% CI: 2.3–4.0), respectively (Figure 1A). 6-month PFS and OS rates were 30.4% (95% CI: 22.5-41.1%) and 54.9% (95% CI: 45.4-66.4%), respectively. 12-month OS rate was 49.5% (95% CI: 38.6 - 63.3%). 29 patients (25%) discontinued therapy prior to completion of one full cycle of treatment with a majority (79.3%) due to progression or death.

**Figure 1.**
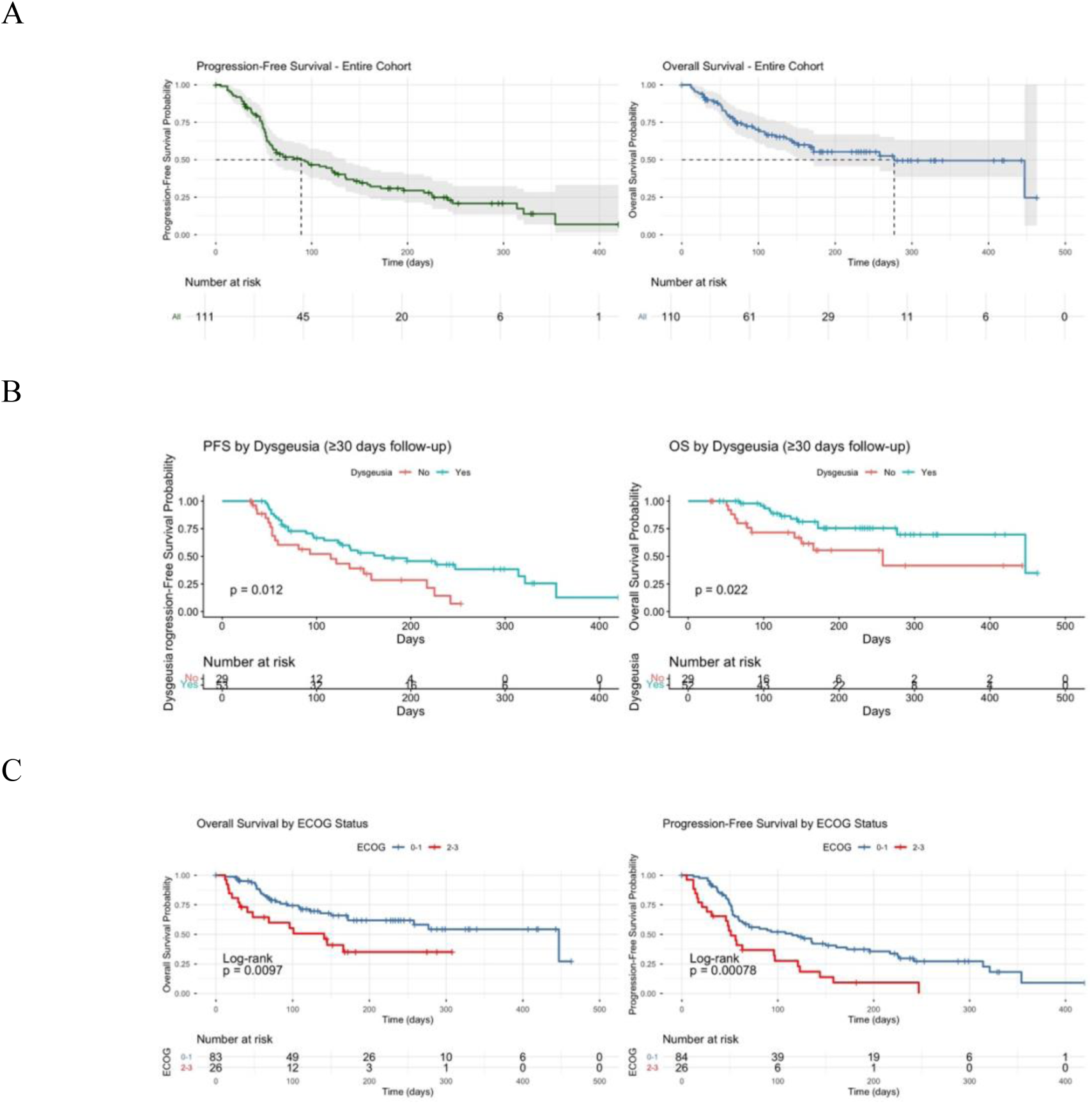
Kaplan-Meier Survival Curves. Patient outcomes on tarlatamab. (*A*) Kaplan-Meier curves for PFS and OS of the entire cohort. Median PFS was 2.9 months (95% CI: 2.3–4.0), and median OS was 9.1 (95% CI: 5.5–NR). *(B)* Kaplan-Meier curves for PFS and OS related to dysgeusia among patients who had received tarlatamab > 30 days. *(C)* Kaplan-Meier curves for PFS and OS related to ECOG at initiation of tarlatamab therapy. CI, confidence interval; NR, not reached; OS, overall survival; PFS, progression-free survival.

### Predictors of Efficacy and Toxicity

In our univariate analysis of baseline biomarkers, several variables demonstrated significant association with grade ≥2 CRS and any-grade ICANS. Most notably, the presence (p=0.035), size (p=0.016), and number (p=0.026) of liver metastases were all associated with the development CRS grade ≥2 (Supplemental Table 1). Likewise, the presence (p=0.007), size (p=0.019), and number (p=0.033) of brain metastases were associated with the development of ICANS of any grade. The presence and severity of CRS were also associated with the development of ICANS (p-0.013 and p=0.027, respectively). Elevated LDH (p<0.001, p<0.001) and metastatic lesion volume outside the liver or brain (p=0.008, p=p0.011) — designated other lesions moving forward — were associated with the development of both CRS and ICANS. Of note, this initial univariate analysis identified significant associations between the presence of a particular variable and toxicity but did not define a threshold level of each variable at which toxicity was associated.

Regarding OS and PFS, our univariate analysis identified increased ECOG performance status, elevated ferritin, and increased disease volume in the brain, liver, and other organs as indicators of earlier progression and increased mortality (Supplemental Table 2). Notably, the presence of dysgeusia was identified as a protective factor associated with improved OS and PFS as well as increased ORR and DCR in patients who had received tarlatamab for at least 30 days.

When corrected for age, sex, and ECOG performance status, our multivariate analysis found that both the sum of liver metastasis diameter ≥10 cm (OR 4.4, p=0.02) and elevated baseline LDH > 250 (p=0.01) were significantly associated with increased likelihood of CRS (Figure 2A). Female sex was a protective factor (OR 0.33, p=0.01). Increased ECOG performance status (OR 2.3, p=0.02), sum of brain metastasis diameter >2 cm (OR 3.6, p=0.02), development of CRS (OR 2.8, p=0.04), elevated baseline LDH > 250 (p=0.04) and elevated baseline ferritin (p=0.004) were all associated with increased risk of developing ICANS (Figure 2B).

**Figure 2.**
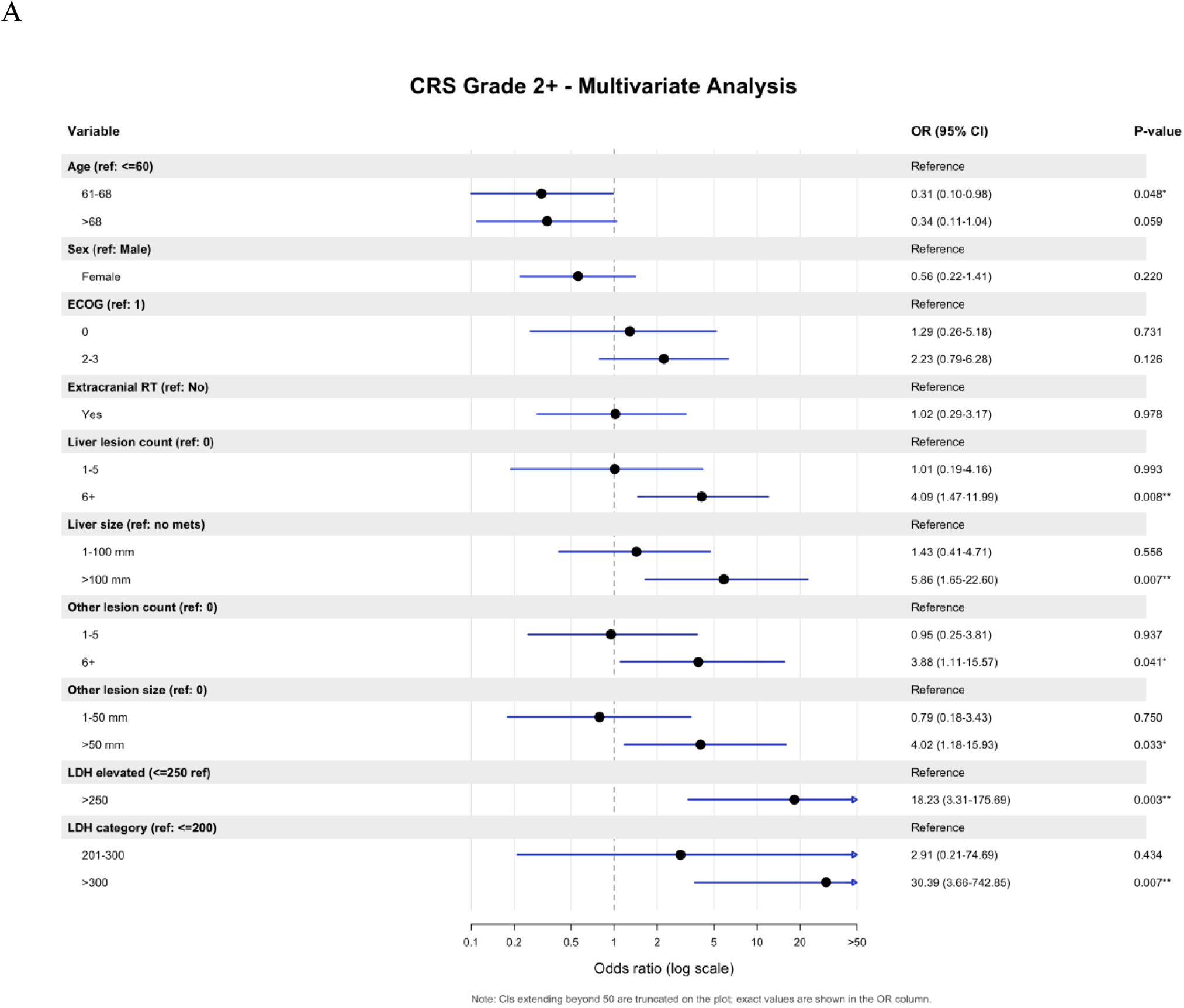
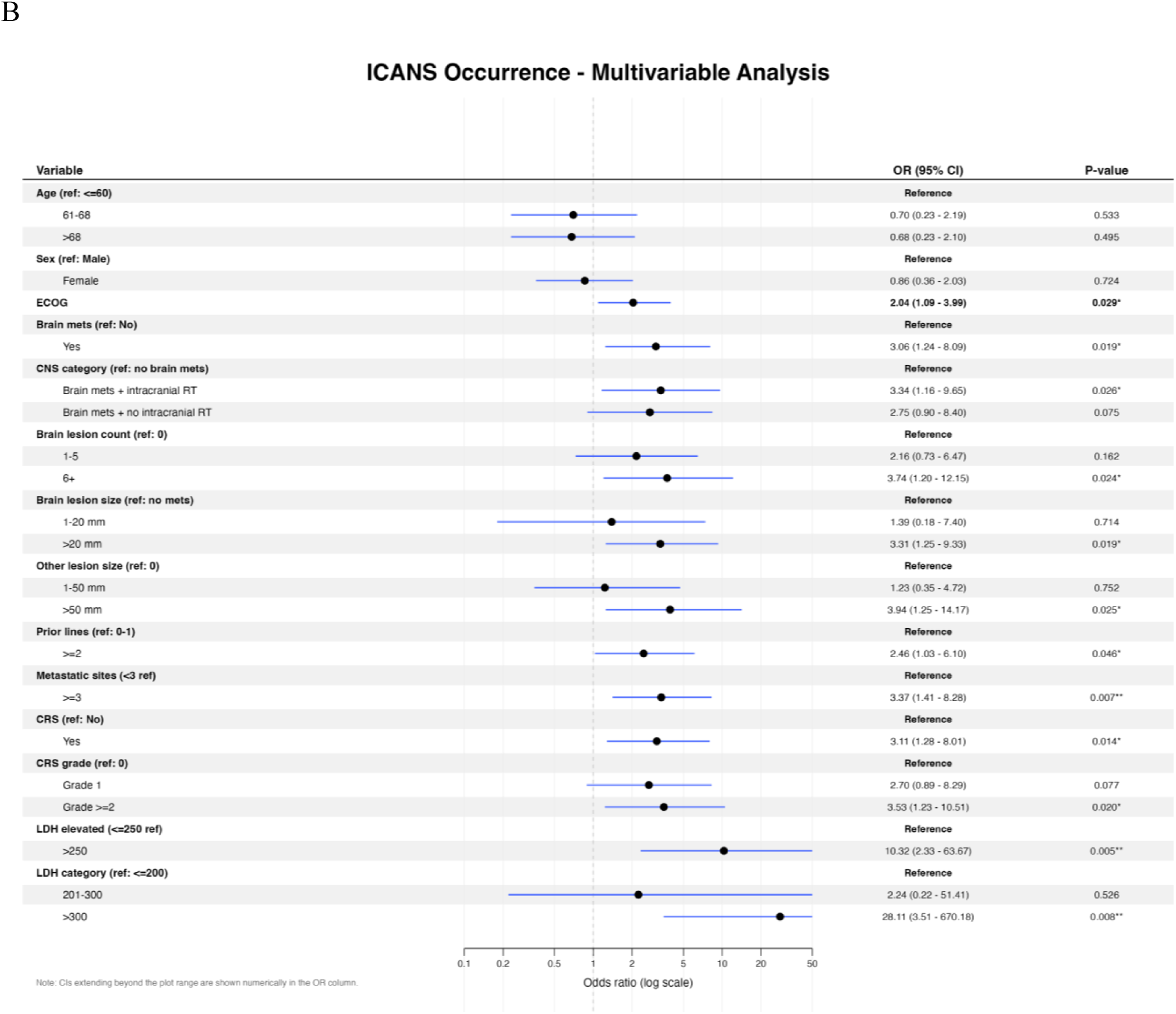
Multivariate Analysis of Biomarkers Associated with CRS and ICANS. Forest plots showing OR and 95% CIs for clinical predictors of CRS grade ≥ 2 and ICANS of any grade. Dots reflect the estimated OR; the dashed horizontal line indicates an OR of 0; the blue horizontal lines represent the 95% CI for each variable. OR, odds ratio; CI, confidence interval; CRS, cytokine release syndrome; ICANS, immune effector cell-associated neurotoxicity syndrome.

Our multivariate analysis of treatment efficacy redemonstrated that the onset of dysgeusia was associated with increased likelihood of disease control (OR 4.26, p= 0.01) and objective response (OR 4.08, p=0.01) (Figure 1B). LDH > 300 U/L, when compared to patients with LDH less than 200 U/L, was associated with increased likelihood of disease control (OR 8.12, p=0.03) and objective response (OR 22.37, p=0.01), while ECOG performance status 2 or 3, when compared to an ECOG of 1, was associated with reduced likelihood of treatment response (OR 0.23, p=0.03) (Figure 3).

**Figure 3.**
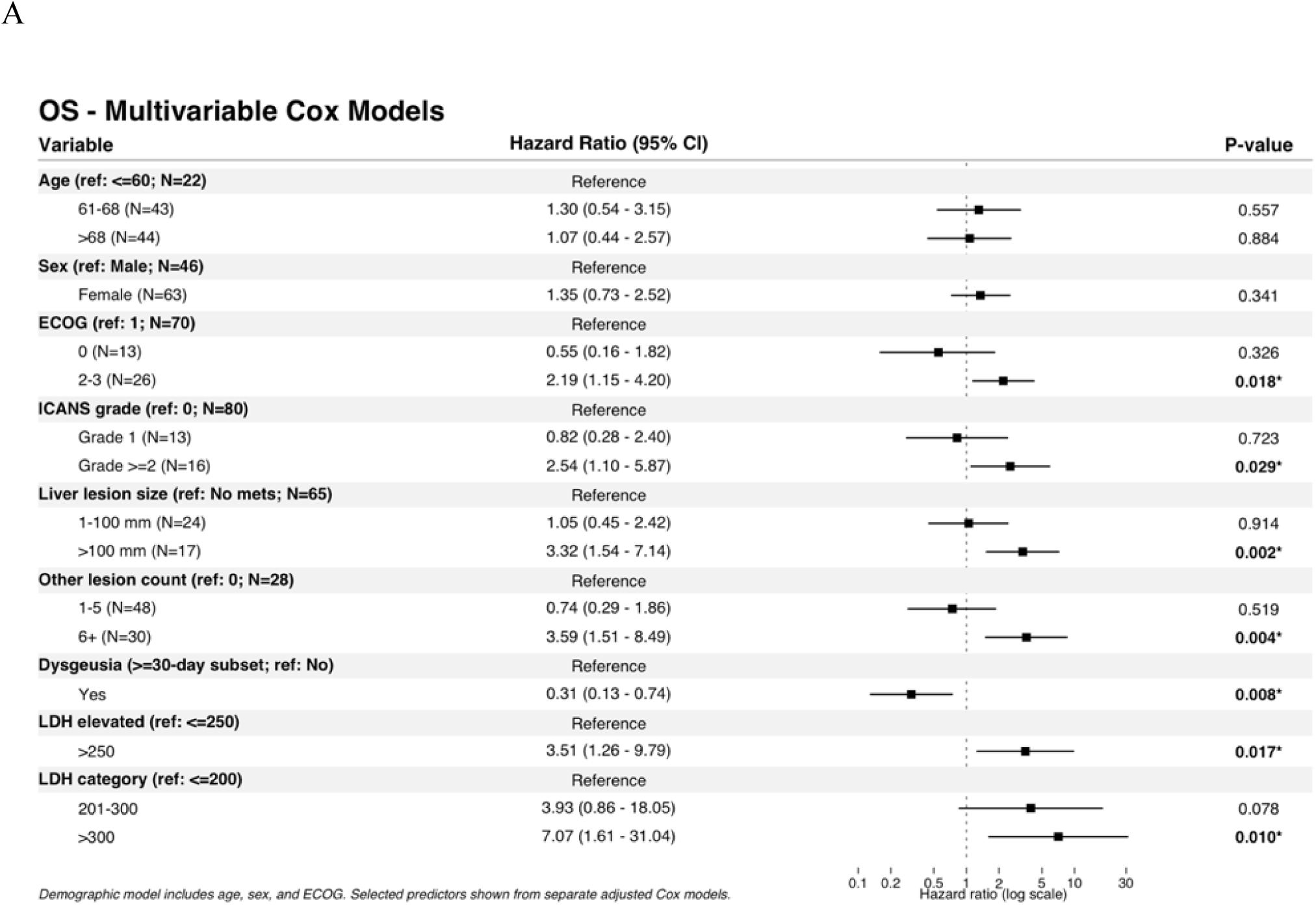
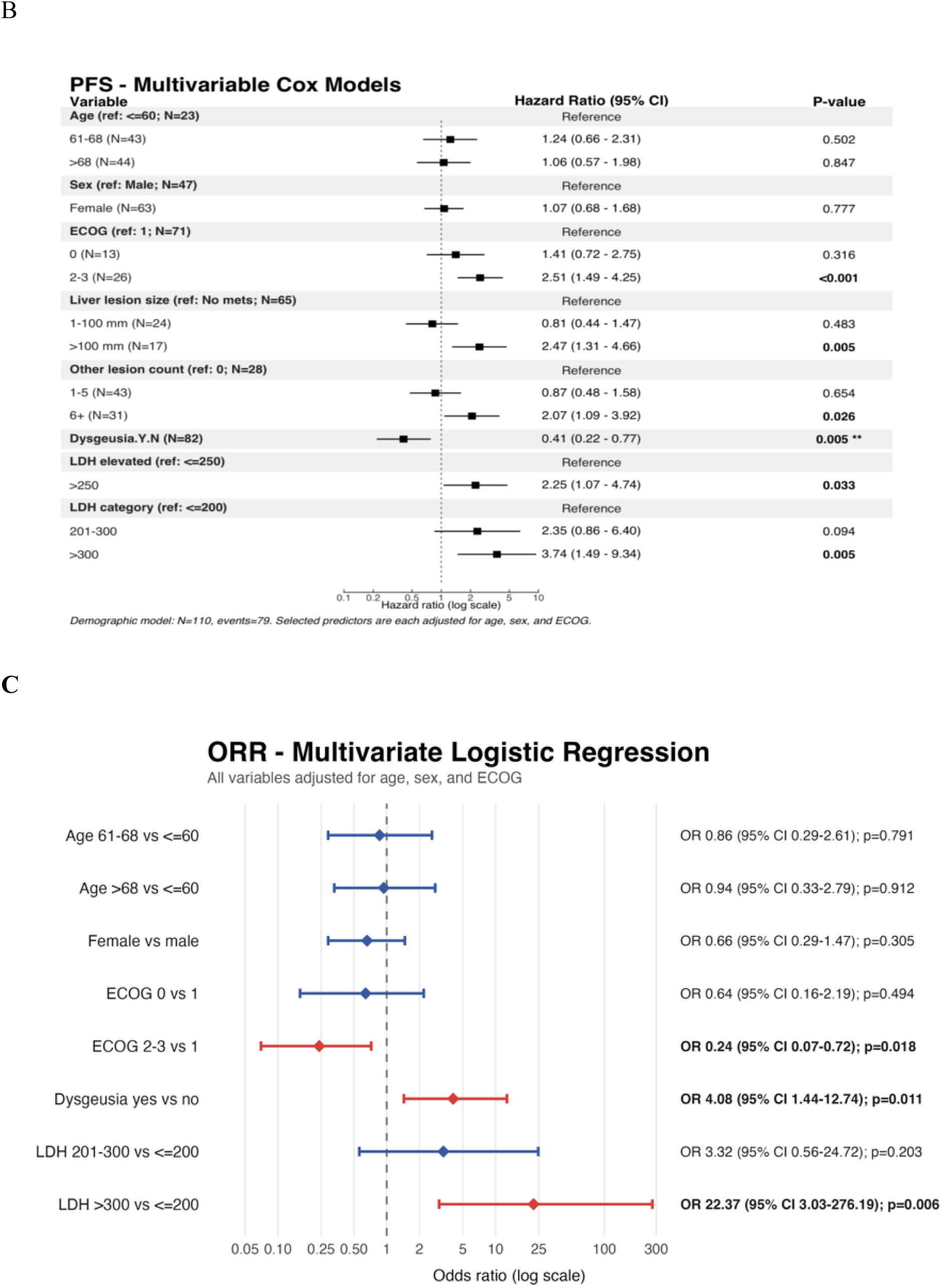
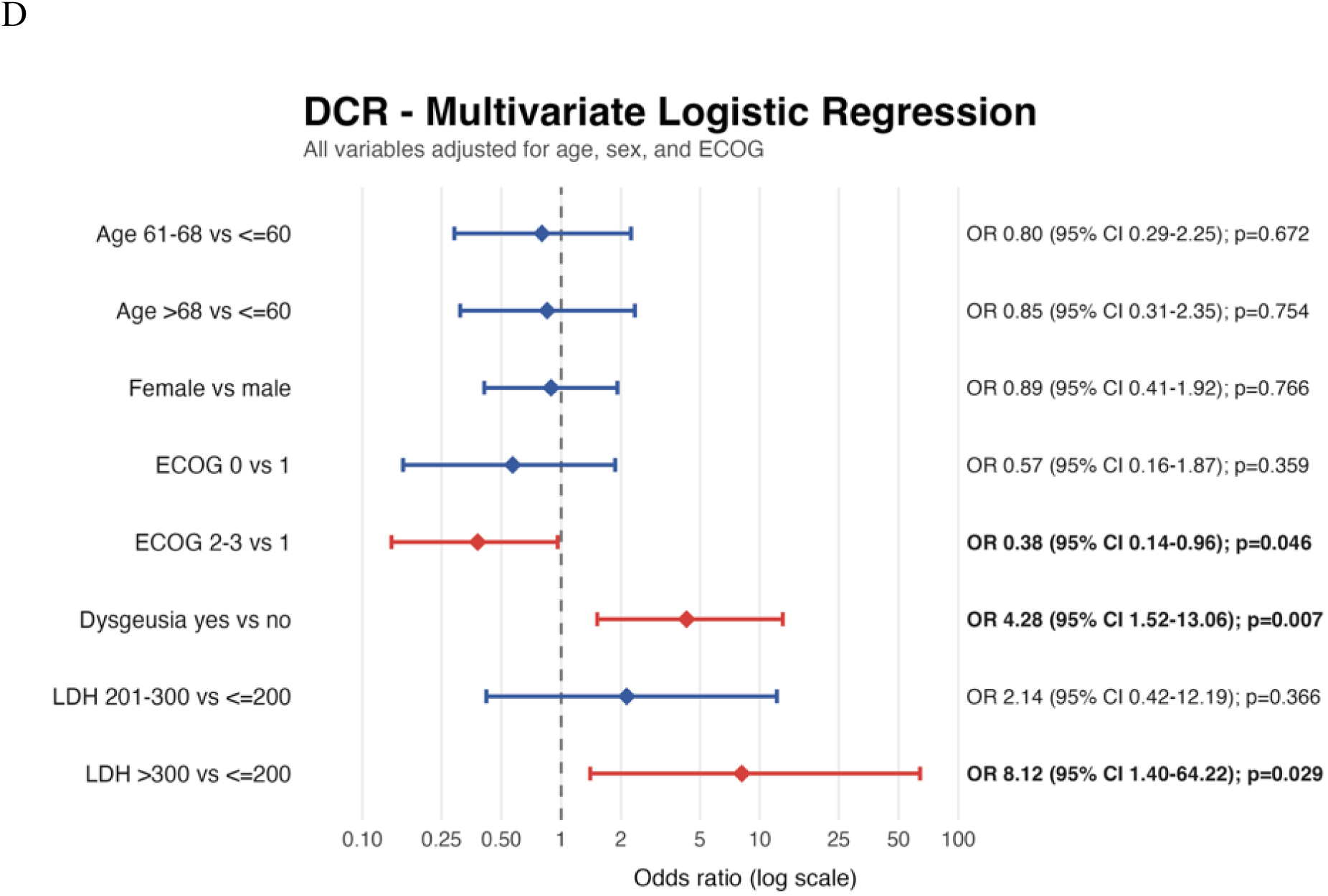
Multivariate Analysis of Treatment Response. Forest plots showing OR and 95% CI for clinical predictors of *(A)* OS, *(B)* PFS, *(C)* ORR, and *(D)* DCR. Dots reflect the estimated OR; the dashed horizontal line indicates an OR of 0; the black horizontal lines in figures 2A and 2B represent the 95% CI for each variable. The red horizontal lines in figures 2C and 2D represent the 95% CI for statistically significant variables. The blue horizontal lines in figures 2C and 2D represent the 95% CI for variables that are not statistically significant. OR, odds ratio; CI, confidence interval; OS, overall survival; PFS, progression free survival; ORR, objective response rate; DCR, disease control rate.

### Risk Stratification Model

In designing our risk stratification methodology, several regression models were considered, with elastic net (EN) ultimately being selected because of its ability to account for correlated predictors while performing coefficient shrinkage and variable selection. In the final EN models, positive coefficients indicated greater predicted toxicity risk, whereas negative coefficients indicated lower predicted risk (Supplemental Table 3). For CRS grade ≥2, elevated C1D1 LDH was the strongest positive predictor (beta=0.396), followed by ≥6 other metastatic lesions (beta=0.295), male sex (beta=0.235), and ≥6 liver lesions (beta=0.189); conversely, 1–5 other lesions (beta=-0.244), female sex (beta=-0.235), and older age (beta=-0.112) were associated with lower predicted CRS risk. For ICANS, ≥3 metastatic sites had the largest coefficient (beta=0.300), followed by worse ECOG performance status (beta=0.265), elevated C1D1 LDH (beta=0.219), and older age (beta=0.085).

The best performing model for CRS grade ≥2 achieved a pooled OOF CV AUC of 0.770, while ICANS prediction AUC was 0.669 (Figure 4A). The CRS prediction model achieved balanced accuracy of >80% in both cross validation and full data, with a sensitivity of >90% when utilizing the full data (Figure 4B). Likewise, the ICANS model had a balanced accuracy of 78% in the full data with a sensitivity of 75%. In the full-data apparent analysis, the CRS model identified nearly all CRS grade ≥2 events, with only one false negative among 24 patients, while the ICANS model had 3 false negatives among 29 patients (Figure 4C).

**Figure 4.**
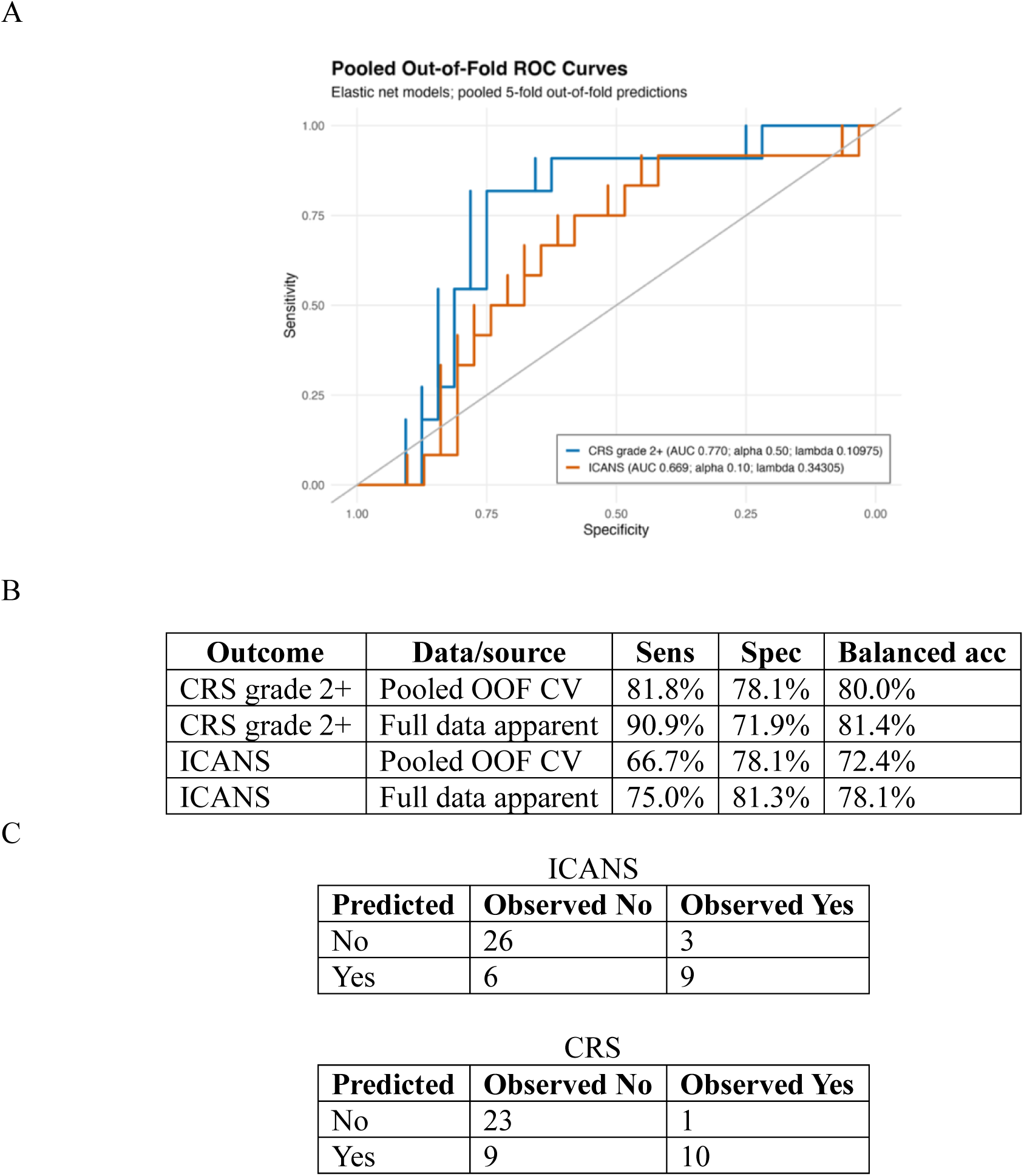
Pooled Out of Fold Cross-Validation (OOF CV) Receiver Operating Characteristic (ROC) Curves for Elastic Net Prediction Model and Associated Performance Metrics. *(A)* ROC curves evaluating the OOF CV AUC for the prediction models assessing for CRS grade ≥ 2 (AUC = 0.770, in blue) and any grade ICANS (AUC = 0.669, in orange). The solid black line represents random chance (AUC = 0.5). *(B)* Performance metrics for pooled OOF CV and full data outlining sensitivity and specificity of each model. *(C)* Confusion matrix utilizing full data apparent after refitting each model on the complete analytic cohort. AUC, area under the ROC curve.

## Discussion

This retrospective, multivariate analysis examined predictive factors associated with tarlatamab treatment response and toxicity. While external validation is required, the statistically derived risk stratification model developed and presented in our results represents an initial tool which demonstrated encouraging ability to predict clinically relevant CRS and ICANS for patients receiving tarlatamab. Together, the results from this exploratory analysis offer clinically valuable insight to assess variables associated with response and toxicity while also providing a preliminary framework to advance future efforts for both escalation and deescalation strategies in monitoring and resource allocation following tarlatamab administration.

Overall, our study population possessed similar baseline characteristics to those of previously reported real-world cohorts; accounting for an increased ECOG performance status within our real-world population, other demographic characteristics such as age and smoking history were similar to clinical trial cohorts(4,5,13). Likewise, the rates of CRS, ICANS, and dysgeusia mirror those of other recent real-world studies, with CRS rates largely in line with those observed in DeLLphi-301(51%) and DeLLphi-304 (56%) trials, however, with notably higher rates of both ICANS and dysgeusia. With an ORR of 44%, our population had largely equivalent response outcomes to those noted in DeLLphi-301 and DeLLphi-304, as well as a comparable signal to other retrospective real-world datasets which have ranged from 34-42.9%(5,6,14). Our cohort demonstrated a diverse spectrum of responses, with 8.8% of patients achieving a clinically noted CR, while 8% of treated patients died within 30 days. This is further reflected in the high early attrition among select patients, with a PFS of 2.9 months across our cohort, which is notably less robust than previously reported clinical trial data, but likely more representative of our sicker, real-world patient population. However, despite early disease progression in several patients, a select cohort demonstrated encouraging, durable responses, as reflected in our 12-month OS rate of 49.5%. Findings from both prospective clinical trial data and available real-world datasets continue to point toward a subcohort of patients who derive enriched and durable benefit from tarlatamab, however, identification of early biomarkers is still needed to better identify and select these patients, particularly as newer systemic therapies with novel therapeutic targets show increasing promise for patients with SCLC (15–17).

Regarding predictors of CRS, Grade ≥2 CRS was associated with an increased volume and number of liver metastases, which has been previously reported in real-world populations(6,18). Importantly, our analysis indicated that this relationship extended to all extrathoracic, extracranial, and non-nodal tumor sites, suggesting an association between CRS and increased tumor burden in general. Likewise, a baseline LDH greater than 250 was associated with the development of CRS. These results are consistent with previously reported data, where elevated LDH and increased tumor burden both generate the release of inflammatory cytokines(19,20). This, coupled with high tumor antigen expression with increased tumor burden, may drive underlying systemic inflammation and predispose patients to the increased risk of CRS(21).

Our ICANS multivariate analysis similarly identified LDH greater than 250 and the presence, size, and volume of intracranial metastases as predictors of neurotoxicity. An increased ECOG performance status and 3 or more metastatic sites were also associated with development of ICANS. While not useful for risk stratification purposes, the development of CRS was interestingly associated with an increased risk for ICANS. Collectively, these findings suggest an observed relationship between select presentations of ICANS, or other acute neurologic changes from tarlatamab, and CRS. As we continue to elucidate the specific immunologic relationship between these toxicities, researchers and clinicians must be careful when labeling various neurologic sequalae from tarlatamab, with other neurologic phenomenon such as tumor inflammation-associated neurotoxicity also described in this setting, suggesting a more nuanced classification may be needed (22).

Strikingly, the only variable associated with an increased likelihood of disease response was dysgeusia. This protective effect was observed once variables were controlled to only include patients who had received at least 30 days of therapy. Decision to exclude patients with < 30 days exposure was purposeful given the known timeline of this particular side effect. Dysgeusia is a well-documented on-target off-site toxicity of tarlatamab that is becoming increasingly common in real-world patients with relevant implications for quality of life (7,23). While these results are intriguing, they must be interpreted cautiously and certainly with an appreciation such side effects have on patients’ ability to tolerate tarlatamab long-term. Importantly, neither CRS nor ICANS was associated with treatment response, reinforcing the complex interplay of immune activation and targeted anti-tumor effect that drives these processes.

The risk models generated using these predictive biomarkers showed high sensitivity in identifying patients at increased risk of developing severe CRS and ICANS. In the CRS model, elevated baseline LDH, male sex, ≥6 liver lesions, and ≥6 other lesions increased predicted risk, whereas older age, female sex, and 1–5 other lesions were associated with decreased risk. The more parsimonious ICANS model retained older age, worse ECOG performance status, elevated baseline LDH, and ≥3 metastatic sites, all of which increased predicted risk. The selection of overall metastatic burden rather than brain metastases for the ICANS model suggests that aggregate disease burden provided greater incremental predictive information for ICANS in this cohort, reinforcing the aforementioned pathophysiologic complexity of neurologic sequalae. While preliminary, the ability of these risk models to consistently identify such high-risk patients using readily available clinical variables is encouraging and has the potential to further optimize the delivery of this therapy.

It is important to note that this analysis was conceived and performed prior to recent changes in FDA regulations regarding tarlatamab monitoring requirements in September 2026, which now specify a monitoring period of 6 to 8 hours with the first two tarlatamab doses. In stratifying individual needs for monitoring and prophylaxis, these models can help facilitate this transition and provide reassurance that patients deemed low-risk can safely initiate treatment with reduced monitoring need. Conversely, the identification of high-risk patients, may also offer more precise employment of inpatient monitoring or inform future prophylaxis strategies. Important to note is the hypothesis-generating nature of this analysis, and the need for prospective validation of our derived risk-stratification model, which is an important future direction of our research.

There are several important limitations to acknowledge in this study. Due to the retrospective, multicenter nature of this analysis, variability in toxicity grading and imaging interpretation may have influenced reported results. Likewise, variation in institutional lab collection practices limited the availability of certain biomarkers for select patients, leading to some degree of missingness within our dataset, potentially impacting both our multivariate analysis and prediction model development. When possible, careful corrections to account for such data missingness was employed. In addition, the study cohort had a notable subpopulation of patients who transitioned to hospice prior to completion of their first cycle, likely skewing survival curves and underestimating long-term toxicities. Finally, while internal validation of the risk models were promising, external validation with a larger, more diverse clinical cohort is necessary to confidently advance this tool into further clinical investigation.

In summary, this exploratory analysis of a real-world cohort of ES-SCLC patients receiving tarlatamab identified several clinical and laboratory biomarkers associated with treatment response and toxicity. A risk stratification tool utilizing selected biomarkers to predict the likelihood of clinically relevant grade ≥2 CRS and ICANS successfully categorized high-risk and low risk populations. These results offer a promising initial framework for future clinical application, with the risk model serving as an easily implementable tool that can help direct healthcare resources and improve the safe administration of tarlatamab. External validation, in larger cohorts, is required to confirm our findings and strengthen the prediction accuracy of this derived model.

## Data Availability

All data produced in the present study are available upon reasonable request to the authors

