## Supplemental Tables for "Predictive markers of response and toxicity to tarlatamab in patients with extensive stage small cell lung cancer: a multi-institutional real-world analysis"

**Supplemental Table 1. Univariate Analysis of Biomarkers Associated with CRS and ICANS.**

| Variable | CRS ≥2 / N by level | Test | P |
|---|---|---|---|
| Liver mets | No: 11/67; Yes: 16/48 | Chi-square | 0.035 |
| Liver lesion count | 0: 11/67; 1-5: 3/16; 6+: 13/31 | Fisher | 0.026 |
| Liver lesion size | No mets: 11/67; 1-100 mm: 6/25; >100 mm: 9/18 | Fisher | 0.016 |
| Other lesion count | 0: 5/29; 1-5: 7/45; 6+: 14/32 | Chi-square | 0.010 |
| Other lesion size | 0: 5/29; 1-50 mm: 5/36; >50 mm: 16/37 | Chi-square | 0.008 |
| LDH elevated >250 | ≤250: 2/30; >250: 11/23 | Chi-square | <0.001 |
| LDH category | ≤200: 1/18; 201-300: 2/16; >300: 10/19 | Fisher | 0.002 |

| Variable | ICANS / N by level | Test | P |
|---|---|---|---|
| Brain mets | No: 9/55; Yes: 23/59 | Chi-square | 0.007 |
| Brain lesion count | 0: 9/55; 1-5: 11/34; 6+: 10/23 | Chi-square | 0.033 |
| Brain lesion size | No mets: 9/55; 1-20 mm: 2/9; >20 mm: 19/47 | Fisher | 0.019 |
| Other lesion size | 0: 5/28; 1-50 mm: 8/36; >50 mm: 18/37 | Chi-square | 0.011 |
| Metastatic sites ≥3 | <3 mets: 15/77; ≥3 mets: 17/37 | Chi-square | 0.003 |
| Prior lines ≥2 | 0-1: 11/58; ≥2: 21/56 | Chi-square | 0.028 |
| LDH elevated >250 | ≤250: 3/29; >250: 12/23 | Chi-square | <0.001 |
| LDH category | ≤200: 1/17; 201-300: 3/16; >300: 11/19 | Fisher | 0.002 |
| Ferritin elevated (>500) | No: 3/23; Yes: 5/10 | Fisher | 0.036 |
| CNS category | No brain mets: 9/55; brain mets + no intracranial RT: 10/27; brain mets + intracranial RT: 13/32 | Chi-square | 0.026 |
| CRS yes/no vs ICANS yes/no | CRS no: 50 no ICANS / 11 ICANS; CRS yes: 32 no ICANS / 21 ICANS | Fisher exact | 0.013 |
| CRS grade 0/1/2+ vs ICANS yes/no | Grade 0: 50/11; Grade 1: 17/9; Grade 2+: 15/12 | Fisher exact | 0.027 |
| CRS grade 2+ vs 0-1 vs ICANS yes/no | CRS 0-1: 67/20; CRS 2+: 15/12 | Fisher exact | 0.048 |

**Supplemental Table 2. Univariate Analysis of Biomarkers Associated with OS/PFS/ORR/DCR.**

**A. OS**

| Variable | Log-rank p |
|---|---|
| Age category | 0.759 |
| Sex | 0.426 |
| ECOG category | 0.024 |
| Ferritin elevated | <0.001 |
| Other lesion count | <0.001 |
| ICANS grade 2+ vs 0-1 | 0.010 |
| Liver lesion size | 0.020 |
| Dysgeusia >=30 day cohort | 0.022 |
| LDH elevated >250 | 0.027 |
| ICANS grade 0/1/2+ | 0.032 |

**B. PFS**

| Variable | Log-rank p |
|---|---|
| Age category | 0.870 |
| Sex | 0.933 |
| ECOG category | 0.002 |
| Ferritin elevated | <0.001 |
| Dysgeusia >=30 day cohort | 0.012 |
| LDH elevated >250 | 0.032 |
| Liver lesion size | 0.041 |
| CNS category | 0.041 |
| Other lesion count | 0.048 |

**C. ORR**

| Variable | P-value |
|---|---|
| Age category | 0.866 |
| Sex | 0.199 |
| ECOG category | 0.036 |
| Dysgeusia>= 30d | 0.033 |
| Other lesion count | 0.027 |
| LDH category | 0.044 |

**D. DCR**

| Variable | P |
|---|---|
| Age category | 0.858 |
| Sex | 0.573 |
| ECOG category | 0.344 |
| Dysgeusia, >=30d cohort | 0.023 |
| Ferritin elevated >500 | 0.050 |

**Supplemental Table 3. Coefficients for final selected prediction model for CRS grade ≥ 2) and any grade ICANS occurrence.**

**A. CRS**

| Term | Coefficient |
|---|---|
| (Intercept) | -1.223 |
| C1D1 LDH | +0.396 |
| Total Other Lesion: 6+ | +0.295 |
| Total Other Lesions: 1-5 | -0.244 |
| Sex - Female | -0.235 |
| Sex - Male | +0.235 |
| Total liver lesion: 6+ | +0.189 |
| Age at C1D1 | -0.112 |

**B. ICANS**

| Term | Coefficient |
|---|---|
| (Intercept) | -1.040 |
| >3 Metastatic Sites | +0.300 |
| ECOG | +0.265 |
| C1D1 LDH | +0.219 |
| Age at C1D1 | +0.085 |
